# A simple numerical and analytical analysis of Covid-19 progression, infection inhibition and control in various countries

**DOI:** 10.1101/2020.08.11.20173203

**Authors:** U. Chakravarty, Deepa Chaturvedi, M.P. Joshi

## Abstract

Covid-19 disease outspread and its subsequent control and inhibition strategies in various countries have been different which led to quite drastic difference in the outcome of the disease progression. In this paper we present an analytical and numerical study of Covid-19 disease spread and control by applying the modified SIR model of epidemic outbreak to explain the Covid spread from February-July 2020 in various countries. Two approaches are evident from the disease progression; one focused on disease eradication and inhibition, and the other is less restrictive dynamic response. Both the approaches are analytically modeled to determine the parameters that characterize the effectiveness of dealing with the disease progression. The model successfully explains the Covid-19 evolution and control of various countries over a vast span of four-five months. The study is highly beneficial to simply analytically and numerically model this complex problem of epidemic proliferation. It assists to easily determine the mathematical parameters for the Covid-19 control measures, helps in predicting the end of the epidemic, and most importantly conceiving the judicious way of unlock process to restore communication between cities, states and countries.

## Introduction

The entire world continues to be severely affected by the global corona-virus pandemic Covid-19 which is a severe acute respiratory syndrome Corona Virus 2 (SARS-COV-2). After the first case was reported in Wuhan China on December 2019, the epidemic has spread rapidly to almost each and every country of the world [1, 2]. In addition to the fatality of the disease, it is also highly communicable in nature and that has put unimaginable stress to the existing health facilities which needs to be upgraded rapidly to cope up with the spread. Since the reproduction number of Covid-19 is high very soon the hospitals of major cities of the world of many countries started becoming overloaded with Covid patients. The mortality from Covid-19 becomes more, from the shortage of ventilator systems or ICU for people with severe respiratory symptoms apart from vulnerable people with precondition. Even before the international flights could be banned or restricted, people be tested or quarantined, many countries started showing Covid patients from the beginning of the year 2020. Due to lack of testing facilities, proper protective kits and equipments and respiratory aids and known medicine, eventually all the countries imposed lockdowns of various degree of severity, promoted social distancing, Covid-19 awareness, encouraged and implemented overall hygiene and hand sanitization [3, 4]. Unprecedented threat to public health, the huge disruptive impact on economy and redefining social interaction norms jeopardizes the overall subsistence and welfare of human kind. While scientists of many countries are fervently trying to find cure and vaccine for the deadly virus, though its discovery, testing, validation, production and distribution to everyone will take many months. Thus every nation is currently managing this disaster using a vigilant approach in adopting, analytical assessment of ongoing situation, strategically formulating ways to save lives as well as finding ways for mitigating its impact on economy. Some countries have been able to bring the situation under control and were able to decrease the infection (China, South Korea, Iran, Turkey etc.) by very strict approach. Some countries were able to control the disease where the number of cases was not allowed to increase after attaining a maximum value (Russia, France, Canada etc).Many countries like USA, India etc. are now trying to contain the disease spread and active cases are still on surge, though recoveries are also showing improvement and in near future control may be expected [5,6].To tackle the disaster it is important to know the disease spreading characteristics like Reproduction number, recovery rate, fatality, symptoms and associated complications, diagnostic and cure. Over the course of few months of the outbreak of Covid-19 a lot of data and information is now known regarding the epidemic. It is important to formulate strategies based on analytical, mathematical and numerical modeling to suggest effcetive lockdown measures [7, 13]. It is also crucial to predict time for infection to peak, maximum infection, and the end of epidemic. Quantifying the government restriction and social awareness and diseases prevention response of society is yet another important requirement [2]. The unlock policies based on the analysis of epidemic progression in various regions, cities, states and countries is also a matter of grave importance. It may be pointed here that a very simple yet effective mathematical model of infection kinetic very well simulated and explained Covid-19 progression in various Chinese provinces [14]. However the simplistic model did not include recoveries, active cases and the model was independent of the population of the region. Therefore in this paper we extended the mathematical formulation of Liang et.al. [13].Using the SIR model (Susceptible, Infected and Recovered) and modifying it to include the total population of the region, recoveries and quantifying the restrictive and preventive measures of the government and people. We present an analytical and numerical study of Covid-19 spread in a local region for different type of approach to prevent the diseases spread. The epidemic prevention can be gradual and dynamic or a rigid preventive measure may be taken. The prior approach is suitable and less restrictive which allows selected movement depending on the present scenario (number of infected people at that moment). This eases economic activity to continue in a dynamically restricted manner. The other approach of absolute rigid measures may be little more effective for control of disease but is economically detrimental. In addition the unlock procedure that is restoring communication between two regions (cities, states or countries) can be easily made based on the quantification of how Covid spread is controlled or inhibited in a region. The regions with similar value or close values of transmission control parameters pose low risk when the movement restrictions are lifted between them. Promoting awareness through, dynamic communication using electronic and print media, smart and agile administration, authentic data sharing and upkeep, collective discipline and adherence to physical distancing and hygiene are instrumental for ensuring the disease control and inhibition and also for preventing mortality.

## The Analytical Model (modified SIR)

We first outline the basic equation of the well know SIR model. Let N_T_ be the total population of a place or a region be it a small zone of outbreak, city, state or a country. Let at time t=t_0_ the initial number of Susceptibles be So, Initial number of Infected be Io and Initial Recovered be R_0_ which is in general zero initially, thus S_0_+I_0_+R_0_=N_T_ (Assuming negligible number of births and deaths during a few months span of time). Similarly instantaneous number of Susceptible S(t), Infected I(t) and Recovered R(t)or (removed) at any time follow S(t)+I(t)+R(t)=N_T_. The three coupled rate equations are outlined

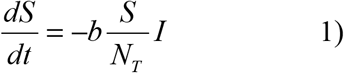

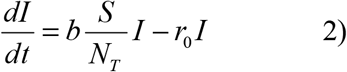

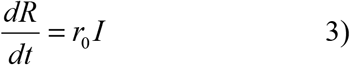

Here b is the transmission rate and r_0_ is the recovery rate. These terminologies erstwhile restricted to books and journals, has now found prominence to a wide audience world over one of them is R_0_ or the reproduction number which depends on the infection rate and recovery rate in a population zone. The normalized quantities s =S/N_T_, i=I/N_T_, r=R/N_T_. We take two forms of b

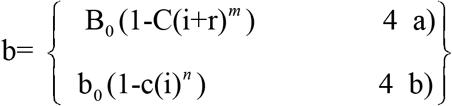

The disease is said to be controlled when active cases start decreasing after attaining a peak value i_m_. The dI/dt value is zero at i_m_, it’s value in general is positive initially during spread and is negative during contol. The fall may be sharp or gradual and in some cases the fall is extremely shallow that the “I” value seems to be almost constant. The epidemic prevention can be rigid and inhibitive depends on the equation 4a) which is more suitable for analysis. Since i+r is the cumulative cases it always increases so “b” value continues to attain smaller values. This approach of absolute rigid measures may be little more effective for control of disease but is economically detrimental due to increasingly stringent restrictions. On the other hand prevention of disease can be gradual and dynamic then equation 4b) is more suitable for analysis. This approach is suitable and less restrictive which allows selective movement depending on the present scenario (number of infected people at that moment). This eases economic activity to continue in a dynamically restricted manner. Majorly the approach used to control the disease is based on the concern of fatalities and over loading of health facilities due to Covid-19 as well as optimizing the economic challenges that the lockdowns and social distancing has brought along. This means the value of i_m_ should be much less that 0.001 (this number depends on available health resources) at any given situation. The whole optimization process makes this restriction of maximum number of cases the top most priority.

## Results and discussion

The Fig. 1 shows the basic outcome for SIR model where time evolution in days is shown for fraction of susceptibles(t), infectedi(t) and recovered r(t) fraction in the total pulation N_T_. The typical values are b_0_=0.5 r_0_=0.1 and a population N_T_ of 1 crore (Here c and C are zero i.e infection control or inhibition is independent of the currently infected or cumulative cases). The number of susceptible decreases sharply after some time and then saturates, similarly the recovered fraction increase rapidly after some days and saturates later. The number of infected (Active cases) increases monotonically first then shows a maximum value of i_m_ (maximum infected fraction) at a time T_p_ and then decreases monotonically. The time where the infected remains only 1% of the i_m_ value may be termed as T_e_(in graph it is shown as T_e_).

**Fig.1.**
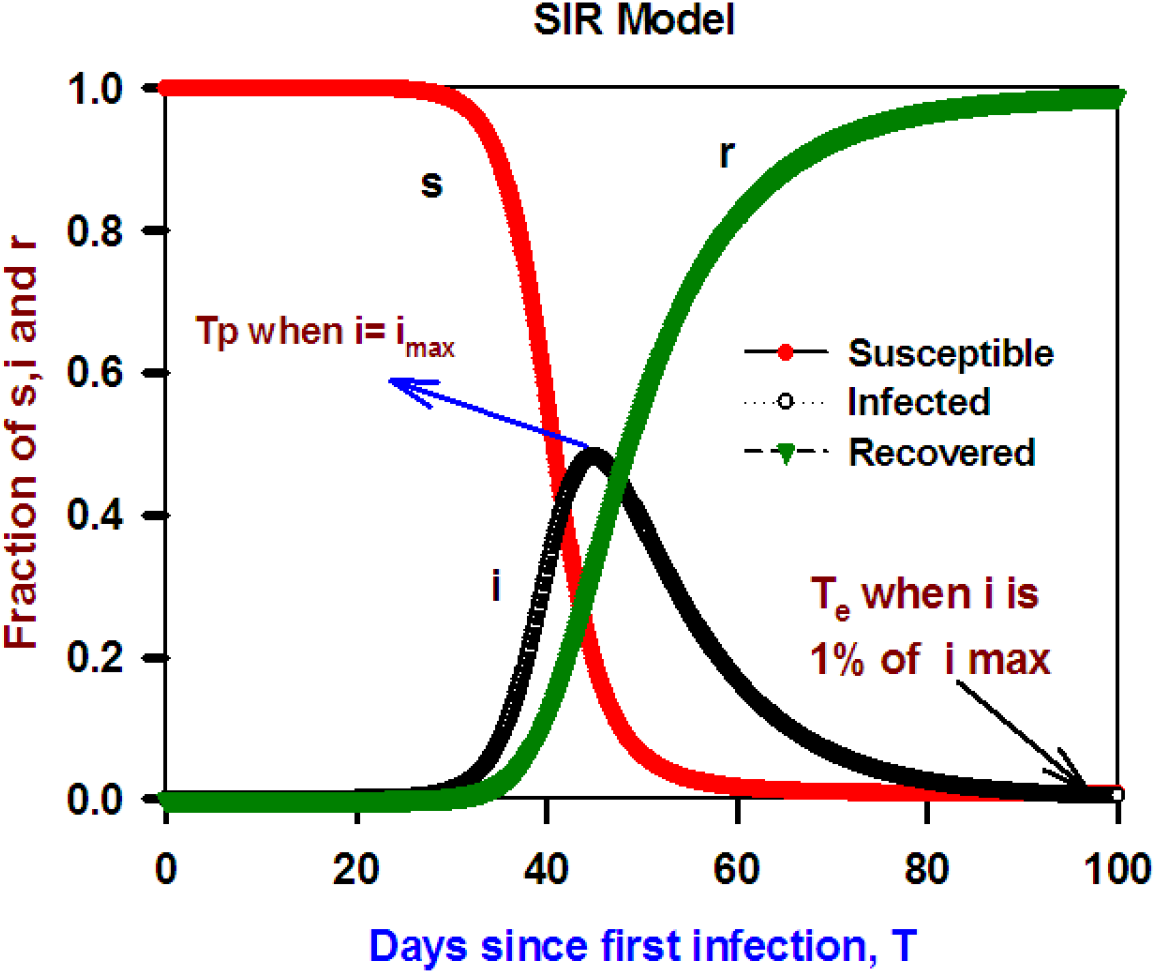
shows the basic curve for SIR model where time evolution in days are shown for fraction of Susceptible s(t), infected i(t) and recovered r(t) fraction in the total population N_T_. The typical values are b_0_=0.5 and r_0_=0.1, and a population N_T_ of 1 crore.

The Fig.2 shows the most important conclusions that can be drawn from the SIR model (using only equation 1 to 3) like predicting the infection peak time, maximum fraction of infected i_m_, and end of epidemic (infected remains only 1% of the i_m_ value) all as function of b (infection transmission rate). The range of values of infection rate is along the X axis. The infection rate of different disease is different highly communicable and infectious have larger b values. “b” can also be changed by implementing, lock downs, social distancing and other restrictive means. The typical value of recovery rate r_0_=0.1 and the population of regions of outbreak of infectious disease are chosen from a small community value to large city population where N_T_=10^3^,10^5^,10^6^of 5*10^6^. This is to bring out the role of total population in disease spread and how the time evolution depends on the population of the region chosen. Here again c and C are taken zero i.e infection control or inhibition is independent of the currently infected or cumulative cases. Fig. 2a) time of infection peaking T_p_ as a function of infection parameter, b. The value of T_p_ monotonically decreases for all population with increasing b. The value of T_p_ is considerably low for lower population region and lower values of b. At high b values and high population T_p_ becomes almost same. Thus it may be concluded that on imposing restrictions and lock down the b value is small (b <1) and T_p_ is much less for low population zones. Thus red containment zones in lockdown is a very effective and fast way to achieve inhibition and control so the disease spread may be contained easily. It is also seen from Fig. 2 b) that the max fraction of infection decreases monotonically on decreasing b (lockdown and social distancing). This is the most important reason for imposing lock downs for a manageable situation ideally i_m_ should be<<0.001. This means only a few people are infected even at peak so that health facility may be available and casualties can be avoided. It may be noted that such a condition can only be obtained by a high degree of awareness by people where they voluntarily maintain social distance, hygiene and remained confined and restricted; only then b will start approaching r_0_ and the disease spread may be inhibited and restricted to a small extent. It is seen that i_m_ is independent of the population of the zone from Fig. 2 b). Thus the disease spreads like a wave and for similar condition of b and r_0_, i_m_ remains same irrespective of the population of the region. Fig. c) shows the time of end of epidemic T_e_ as a function of infection parameter b. The value of Te monotonically decreases for all population with increasing b. The value of Te is considerably low for lower population region and lower values of b. Again high b values and high population Te becomes almost same. Thus it may be concluded that on imposing restrictions and lock down the b value is small (b <1) T_p_ and T_e_ is much less for low population zones. Thus red containment zones in lockdowns are a very effective and fast way to eradicate the disease.

**Fig.2.**
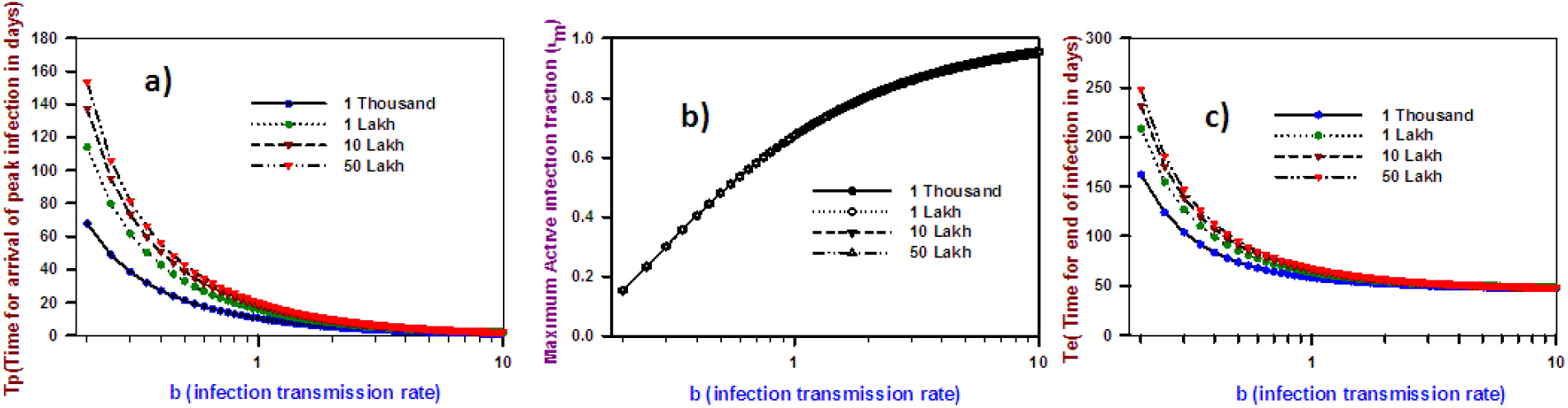
shows the major inferences that can be drawn from the SIR model like predicting the infection peak time, maximum fraction of infected i_m_, and end of epidemic. The population of regions of outbreak of infectious disease is chosen from a small community value to large city population. Fig. 2a) time of infection peaking T_p_ as a function of infection parameter b. Fig. 2 b) max fraction of infection im decreases monotonically on decreasing band is independent of the population Fig. c) time of end of epidemic Tend as a function of infection parameter b.

### A) The countries where the form of disease inhibition followed is B=B0-C(i+r)^m^

#### 1) China

The Fig. 3) The progress of Covid-19 Active cases in China since Jan 22, 2020 when 554 cases were recorded [2,13]. The active case or infection fraction are plotted with days (up to 140 days) peaked at 26 days. The outcome implies extremely strict government action and high degree of awareness amongst the people due to high value of C=1250. The reported data is well simulated by the analytical and numerical model. The analytical form of infection transmission parameter B=B_0_-C(i+r)^m^, here m=1. As evident a highly restrictive disease control regimen was implemented and followed for a period of nearly 5 months. The dynamics is easily modeled by simple analytical forms over a very long period of time and is testimony to the fact the action of government and people have followed the same behavior over a prolonged period of time.

**Fig. 3).**
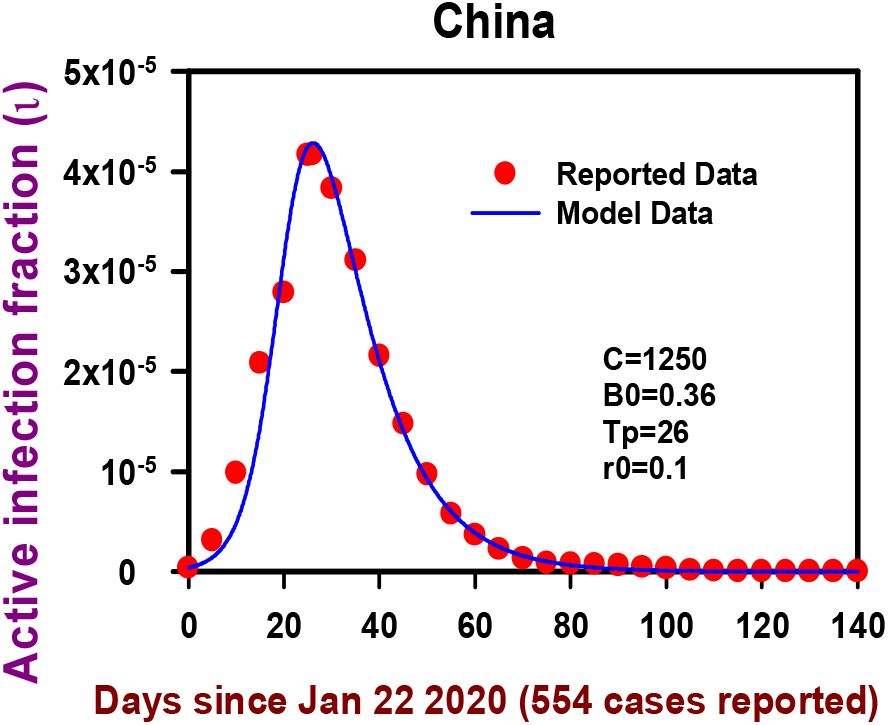
Covid-19 Active cases in China cases since Jan 22, 2020 both reported data and model data show good coincidence.

#### 2) Turkey

Fig. 4) The progression of Covid-19 Active cases time line in Turkey since March10, 2020 when the first case was recorded [14]. The active case or infection fraction are plotted with days (up to 140 days) peaked at 42 days. The outcome implies a strict government action and considerable awareness amongst the people. The reported data is well simulated by the analytical and numerical model. The analytical form of infection transmission parameter B=B_0_-C(i+r)^m^, here m=0.125. A vigilant restrictive disease control regimen was implemented and followed for a period of more than two months. The deviation of reported data from the model beyond 70 days clearly shows a lifting of restrictions and therefore the chosen form of infection has to be modified. Yet again the epidemic dynamics is easily modeled by simple analytical forms over a substantial period of time.

**Fig. 4).**
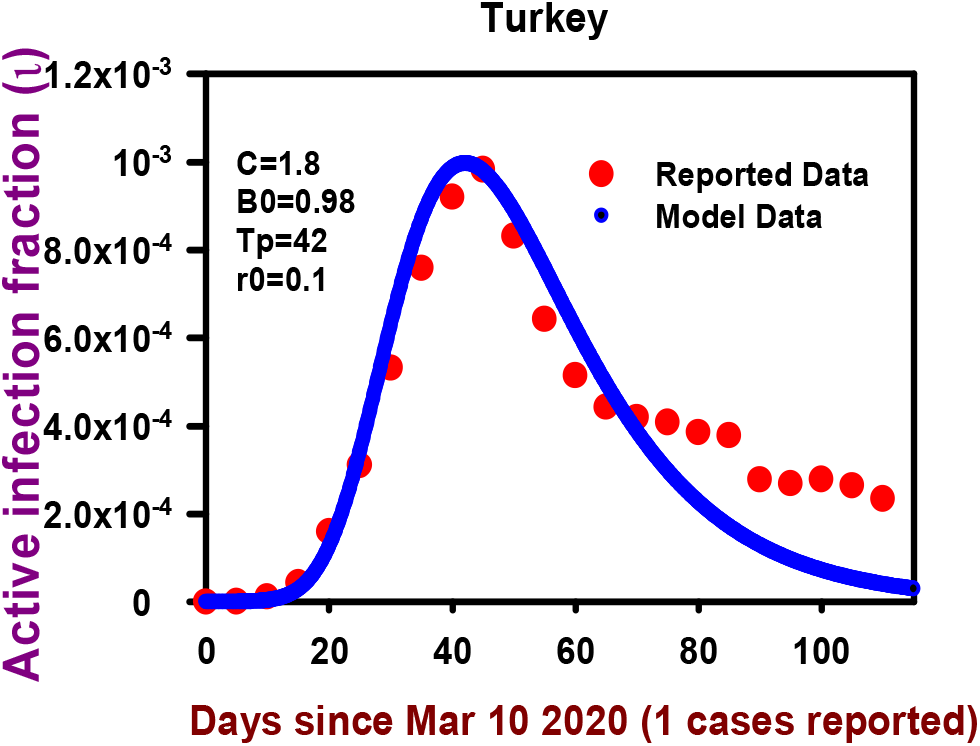
The progression of Covid-19 Active cases time line in Turkey since March 10, 2020. Reported data and model data are close over a long duration of time.

#### 3) Italy

Fig. 5) the progression of Covid-19 Active cases time line in Italy since Feb15, 2020 when 3 cases were reported. The active case or infection fraction are plotted with days (up to 140 days) peaked at 66 days. The actual data shows a steep rise in the infection and control measures seems to be inadequate. Infact after the surge of Covid cases the government of Italy went in to lock down on March 9, 2020 [3]. Since then gradually the strict government action combined with considerable awareness amongst the people the disease was brought under control after peaking at 66 days. It is therefore quite clear that the reported data is well simulated by the analytical and numerical model only after some time, when the extreme measures were implemented. The analytical form of infection transmission parameter is B=B_0_-C(i+r)^m^, where m=0.125. A vigilant restrictive disease control regimen was implemented as well as people adopted social distancing norms which followed for a period of about 3 months. The coincidence of reported data from the model from 50-140 days after the outbreak clearly shows that the epidemic dynamics is easily modeled by the above mentioned simple analytical form.

**Fig. 5).**
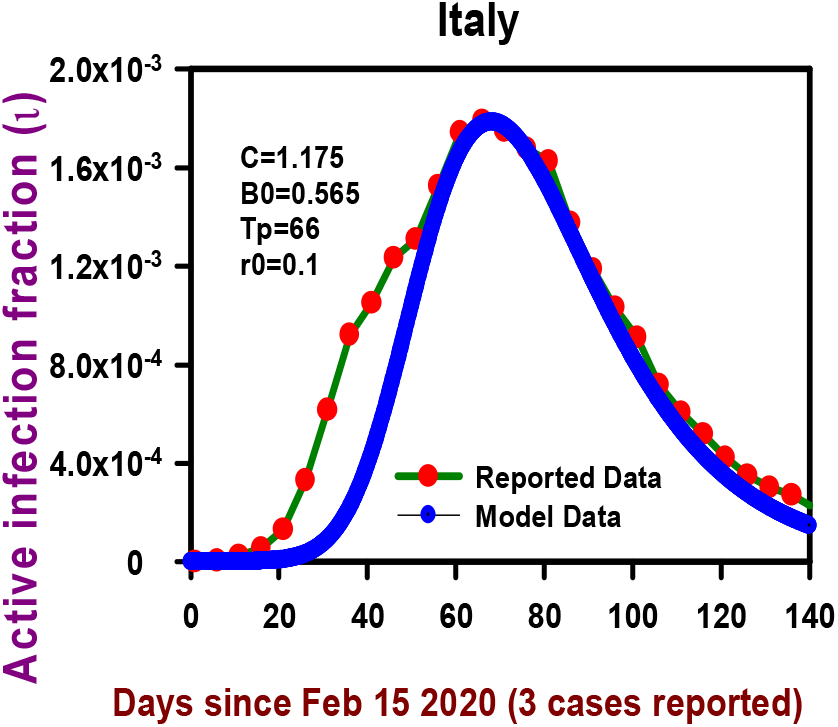
The progression of Covid-19 Active cases time line in Italy. Reported data and model data are close after the implementation of strict disease inhibitive measures.

### B) The countries where this form of disease inhibition is followed b=b_0_-c(i)^n^

#### 1) Russia

Fig. 6) The progression of Covid-19 Active cases in Russia since March 04, 2020 when the first case was reported. The active case or infection fraction is plotted with days (up to 125days) increased rapidly and started to get steady. The infections graduallymaximized at 106 days and became steady thereafter. The outcome implies moderately strict government action and high degree of awareness and a smart and dynamical response amongst the people [15]. The reported data is well simulated by the analytical and numerical model. The analytical form of infection transmission parameter b=b0-c(i)^n^, here n=0.5. Clearly the mode appears that depending on the active cases the response was affected, an increase in infected people was followed by very strict government action and public social distancing.Whereas on the decrease of infection restriction was judiciously loosened. The dynamics is easily modeled by simple analytical forms over a four month long period of time.

**Fig. 6).**
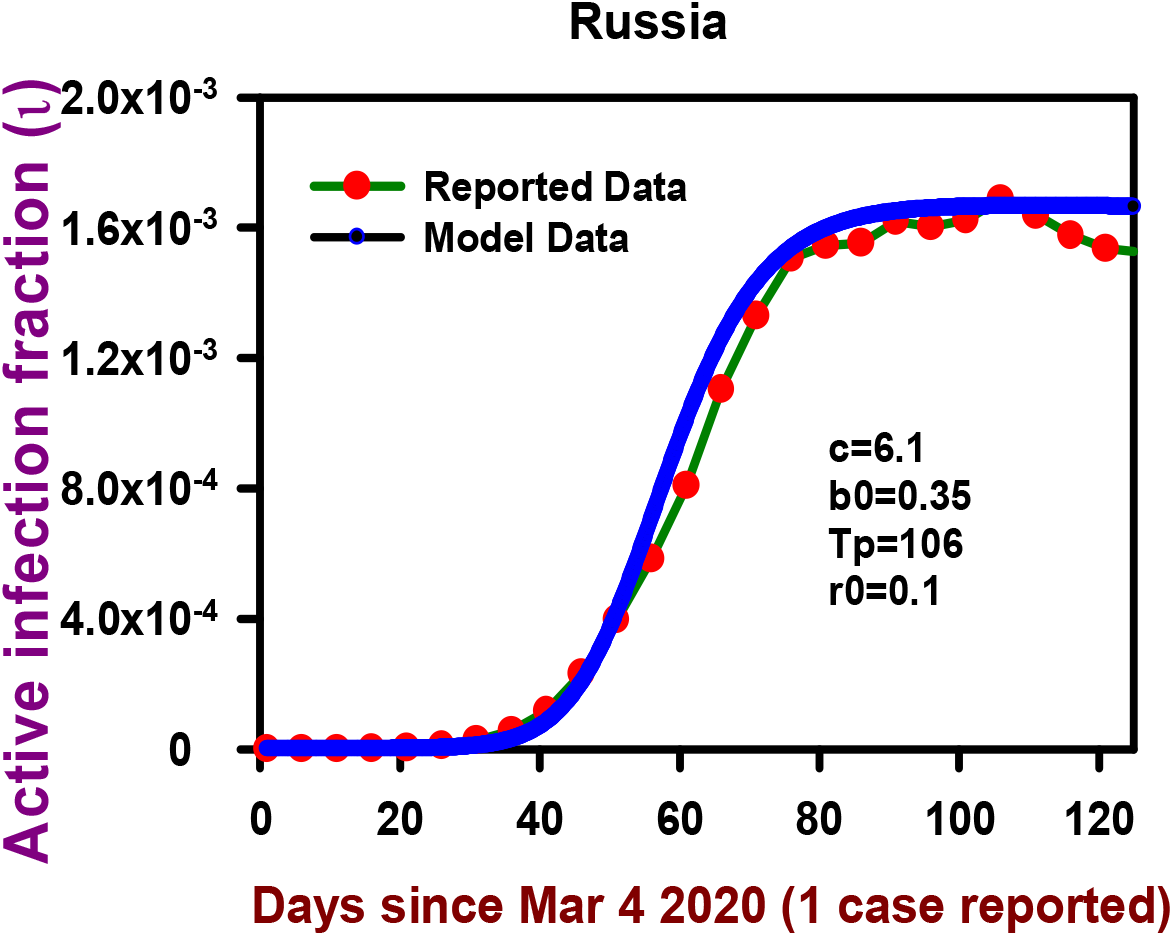
The progression of Covid-19 Active cases in Russia active cases saturated instead of going down indicating a comparatively mild restrictive approach. Choosing b=b0-c(i)^n^, here n=0.5 reported data and model data are very close.

#### 2) France

The Fig. 7) shows the spread of Covid-19 Active cases in France since Feb15, 2020 when the 7 cases were reported. The active case or infection fraction is plotted with days (up to 140days) infection increased rapidly initially. Seeing this lock down period was implemented from 17 March, 2020 to 11 May 2020 and the outcome show it was a progressive and controlled lock down. Control not elimination was the route chosen, infection eventually started to get steady [3]. The infections gradually maximized at 58 days and became steady thereafter. Similar to Russia the outcome implies moderately strict government action and high degree of awareness and a smart and dynamical response amongst the people. The reported data is well simulated by the analytical and numerical model. The analytical form of infection transmission parameter b=b0-c(i)^n^, here n=1.0. Here too, the response (government and people) depended on the active cases at any given instant of time. The dynamics is easily modeled by simple analytical form over a four month long period of time.

**Fig. 7).**
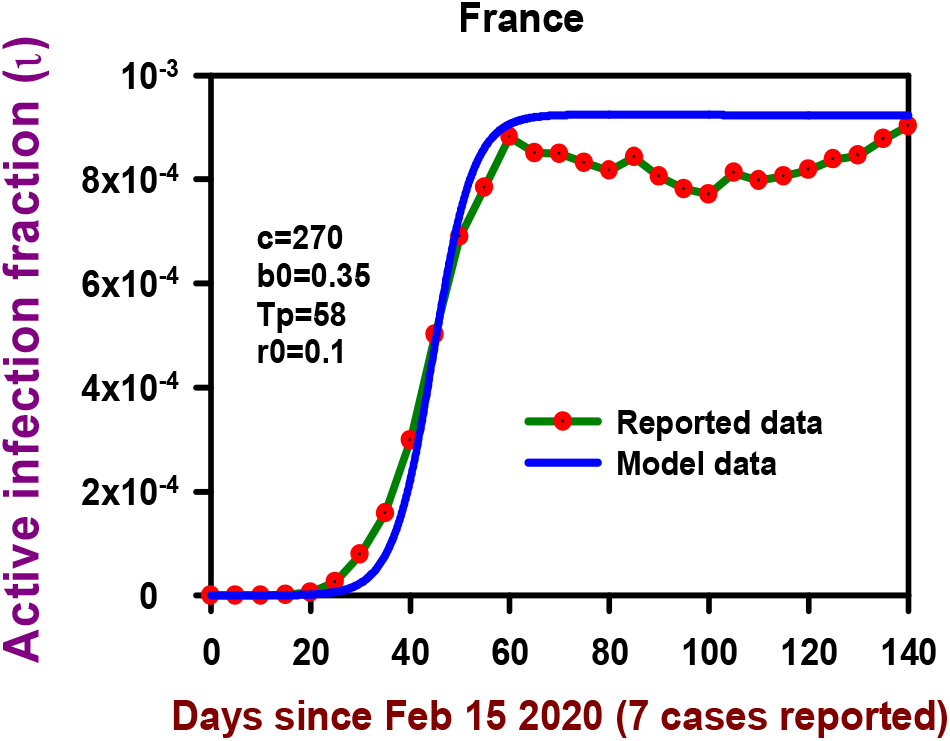
The progression of Covid-19 Active cases in France since Feb 15 2020 when the 7 cases were reported. Choosing b=b0-c(i)^n^, here n=1, like Russian data (Fig.6) reported data and model data are very close.

### Predicting the time peak/ maximum, end of epidemic and maximum infection using modified SIR model

It is important to know the effect of lockdowns and social distancing on peak time of infection, end of epidemic and maximum fraction of infected people. The Fig.8) shows a summary of effectiveness of infection inhibition parameter C. Peak time of infection, end of epidemic and maximum fraction of infected people are calculated from the modified SIR model with the modified infection transmission is taken of the form B=B_0_-C(i+r) which depends on the cumulative cases (active cases+ recovered cases). This form is a restrictive and inhibitive form of response combining both government and public response. The population chosen is 1 crore and typical values of B_0_=0.4 and r_0_=0.1 from the general available Covid progression data. The peak time Tp slightly decreases on increasing C (infection inhibition) thereafter almost becomes constant. This clearly shows no matter how large C is chosen it has no impact on the Tp. Similarly Te gradually and very slowly decrease with increasing C. This again shows the end of epidemic time cannot be brought by more and and more inhibition and restriction, that is increasing C does not have much impact on ending the epidemic. Yet it may be pointed C is very crucial to control the maximum infected fraction i_m_. An increase in C causes a sharp decrease in i_m_ and then gradually the decrease in i_m_ becomes less. Therefore the increasing C beyond a certain degree is not very beneficial. Since a very high value of C is detrimental for the minimum social interaction for economic and livelihood activities, it does not affect T_p_ and T_e_ and i_m_ does not get significantly reduced. Ideally i_m_ less than 10^-3^(depending on health care resources of the region) would be manageable to provide hospitalization for ventilators or ICU.

**Fig.8).**
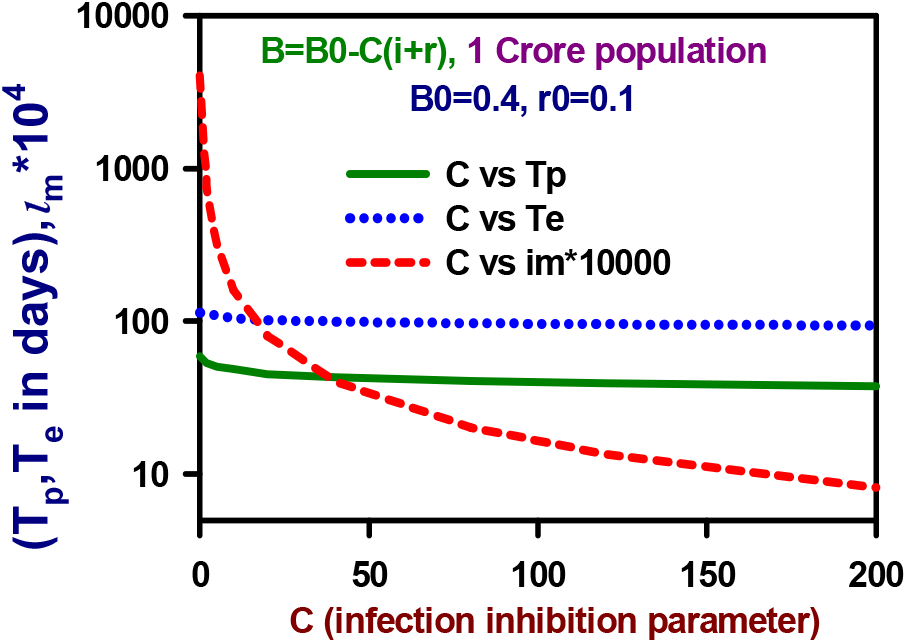
Peak time of infection, end of epidemic and maximum fraction of infected people are calculated from the modified SIR model with the modified infection transmission is taken of the form B=B_0_-C(i+r) which depends on the cumulative cases (active cases+ recovered cases).

Next, the Fig.9) shows a summary of effectiveness of infection control parameter c. Time of maximum infection, end of epidemic and maximum fraction of infected people are calculated from the modified SIR model with the modified infection transmission taken of the form b=b_0_-c(i) which depends on the active cases. As stated earlier this form is less restrictive and mildly inhibitive. “The so called living with disease mode” form of response combining both government and public response is better described by the above form of infection transmission parameter. The population chosen is 1 crore and typical values of b_0_=0.4 and r_0_=0.1. The maximum infection time T_m_ almost constant and does not vary much with the increasing value of c, the infection control parameter. This clearly shows no matter how large C is chosen it has no impact on the T_m_. An increase in C causes a sharp decrease in i_m_ and then gradually the decrease in i_m_ becomes less. Therefore the increasing C beyond aa certain degree is not very beneficial. As mentioned earlier very high value of C is detrimental for economic activities. The value of c must be chosen such that i_m_ much less than 10^-3^ (depending on health care resources of the region). So social activities may be allowed restrictively while minimizing the life threatening outcome to the best possible extent. Similarly in contrast to Fig.8 Te gradually and monotonically increases with increasing c implying disease stay for a long time. Eventually C is very crucial to control the maximum infected fraction i_m_.

**Fig.9).**
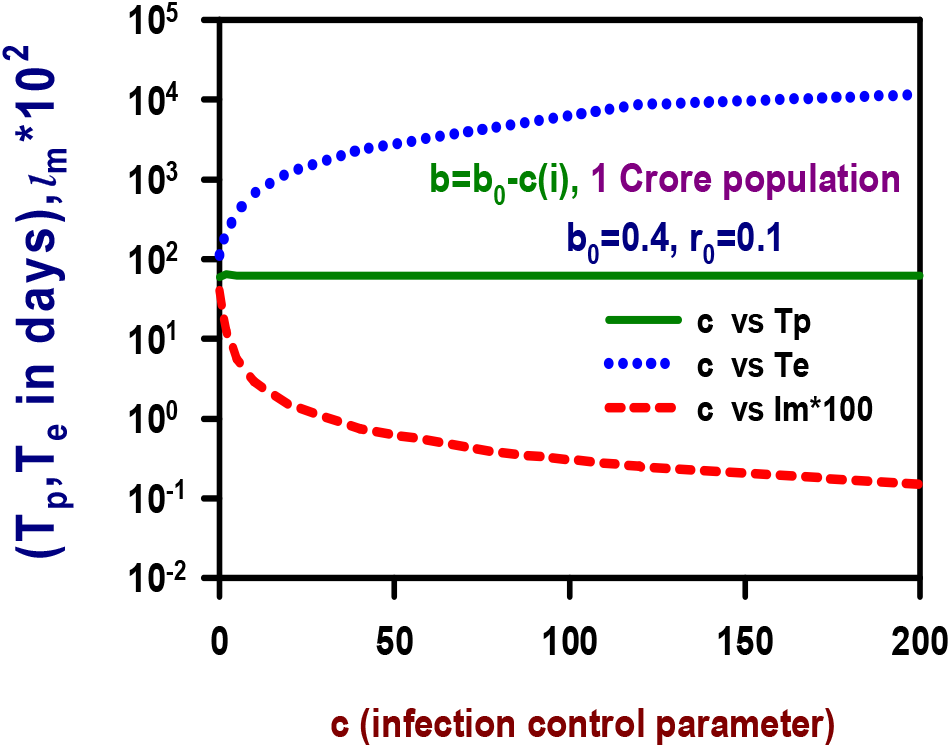
Time of maximum infection, end of epidemic and maximum fraction of infected people are calculated from the modified SIR model with the modified infection transmission taken of the form b=b_0_-c(i) which depends on the active cases.

### Determining the Value of B_0_ and C of various countries assuming B=B_0_-C(i+r)

**Table I:**
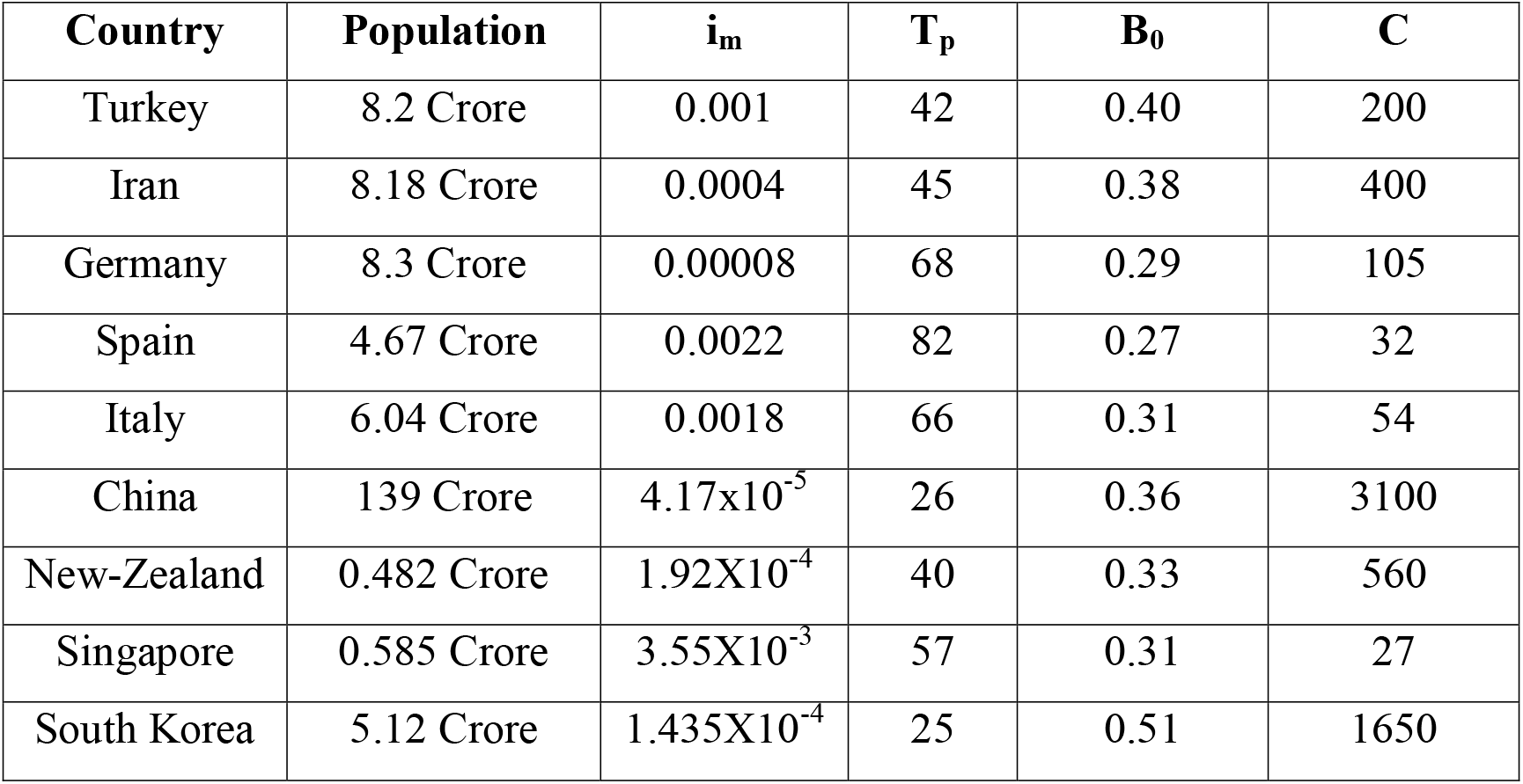
The evaluation of the parameters associated with modified SIR model with B=B_0_-C(i+r) the Value of B_0_ and C of various countries are obtained by matching reported Tp and i_m_ values. The values are also calculated for Iran, Germany, New-Zealand and South korea as these countries applied a strict disease inhibition regime [16,19]. The chosen form of B is based on the observation that the Covid spread was controlled and inhibited and there was a decrease in Active cases after the peak. Note in these cases m=1 the exact matching of reported Tp and i_m_ values is ensured but the trend of reported data and model match only partially.

### Determining the Value of b_0_ and c of various countries assuming b=b_0_-c(i)

**Table II:**
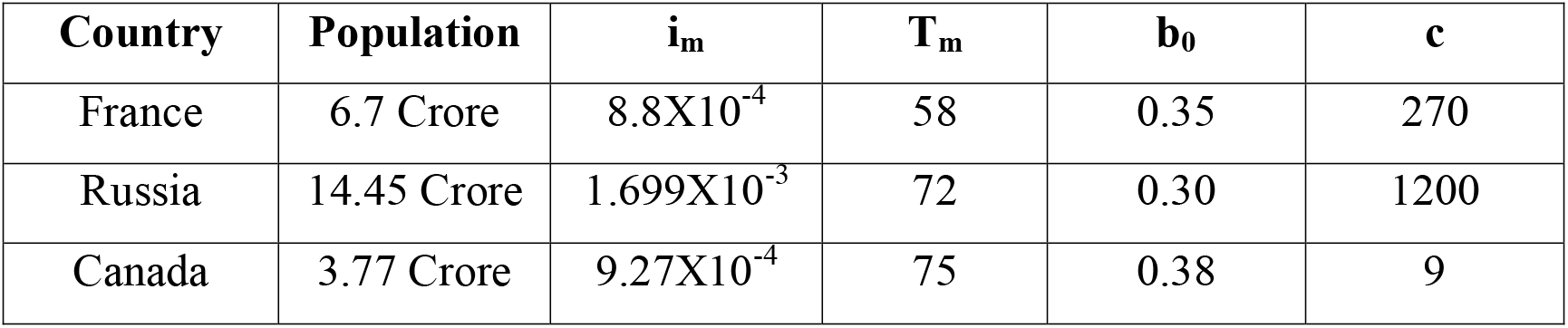
The evaluation of the parameters associated with modified SIR model with b=b_0_-c(i) the Value of b_0_ and c of countries like France, Russia and Canada [20] are obtained by matching reported T_m_ and i_m_ values. The chosen form of b is based on the observation that the Covid spread was controlled in a more gentle manner where Active cases flattened after the peak. Yet again it may be noted in these cases n=1 the exact matching of reported T_m_ and i_m_ values is ensured but the trend of reported data and model match only partially.

### Using the modified SIR model for prediction of Covid-19 progression in India

At last we do a bit of predictive analysis for Covid-19 progression in India. Similar studies have been done in great detail using different analytical models for India and its constituent states [21-23] Various countries like USA, India, Brazil etc. are witnessing a surge of Covid infected patients every day and the disease peak has not yet been reached. India’s fight against Covid is being eyed closely since it is a country with a large population of 135 crore and it also has huge population density. Thus Covid poses a great threat to health of Indian people as well as has repercussions for a setback to the economy. However, the Indian government has implemented timely action through a strict lock down witnessing closure of transport networks, public gatherings, religious places, Schools, Restaurants etc. There were various phases of Lock down 1.0, 2.0, 3.0 and 4.0 starting from 25 March 2020 -31 May 2020. The unlock process started mildly and gradually from June 1, 2020 and unlocking is continuing with various restrictions. In such a dynamic scenario the prediction of disease progression is difficult yet a few inferences can be made from the available data till date where both the government and public social distance response is estimated. Based on which the arrival of peak and the maximum number of people expected to be infected can be predicted. The Figure 10 shows the spread of Covid-19 in India since March 03, 2020 when 3 cases were reported. The reported data available till July 23, 2020 is fitted with modified SIR model with B=B_0_-C(i+r)^m^,with B_0_=0.25, C=0.26 and m=0.125. The lower figure is a zoomed version of the above figure. The peak is expected near October end of 2020. The effect of unlock and overlooking social distancing can have a highly damaging consequence to the people of India. If B_0_ is increased to 0.3 while keeping C=0.26 the maximum number of infected at peak are more than 1%. If B_0_ is kept at 0.25 and C reduced to 0.2 the peak may be expected in November end, with about 1% of population being infected at peak. If both government and people relax the disease inhibition measures by increasing B_0_ to 0.3 and decreasing C simultaneously to 0.2, the situation will become worse and 5% people may be affected. Therefore it can be concluded that the commendable effort of government and vigilant approach of people of India till now, must be continued at all expense to keep the situation under control.

**Fig.10).**
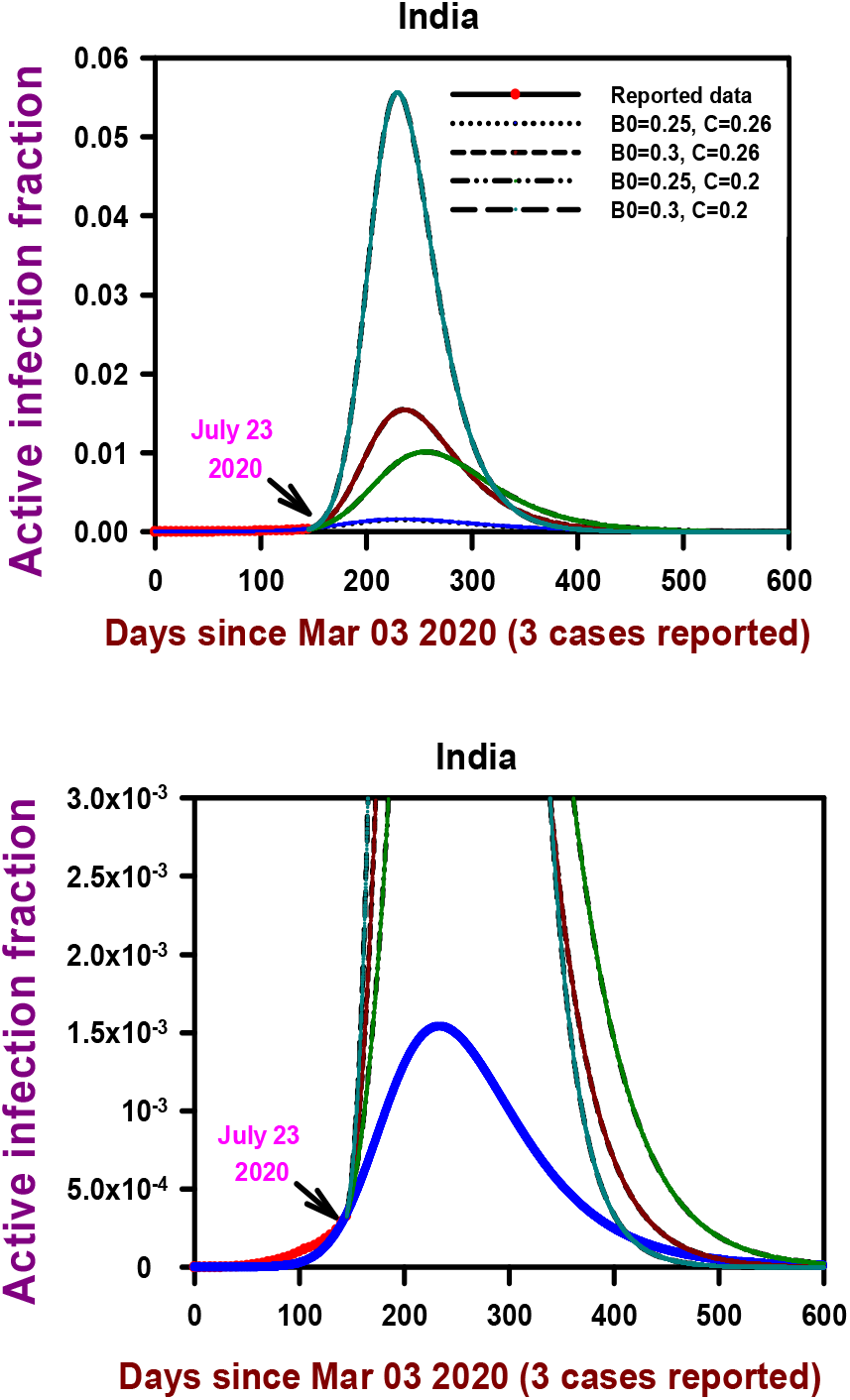
The Covid-19 progression in India since March 03, 2020 when 3 cases were reported. The reported data available till July 23, 2020 is fitted with modified SIR model with B=B_0_- C(i+r)^m^,with B_0_=0.25 and C=0.26. The lower figure is a zoomed version of the above figure. The peak is expected near October end of 2020. The effect of unlock and overlooking social distancing can have a highly damaging consequence to the people of India. The commendable effort of government and people must be continued to keep situation under control.

## Conclusions

In conclusion following points are outlined which summarizes the major results and inferences of the analysis presented in this paper based on reported Covid-19 data.

1. The modified SIR model used in this work explains the time for infection to peak, maximum infection, the end of epidemic as well as helps in quantifying the government restriction and social awareness and disease prevention response of society.
2. The Covid-19 epidemic proliferation dynamics is modeled using analytical and numerical means using two major forms of time dependent infection rate. In first scenario a strict restrictive regimen is imposed for disease inhibition and eradication (China, Italy, Turkey) and in the second control is achieved in a more relaxed manner where the active cases are not let to grow any further after reaching a maximum value (France, Russia,Canada). The first approach is a lot more challenging and taxing for the economy and social activities. The other approach is more relaxed where a smart and dynamic way is adopted by the government and people to safeguard economy while ensuring the control of disease spread. Both these approaches are numero-analytically modeled and explained in detail the reported Covid-19 data recorded over a period of 5 months.
3. The model clearly brings out the role of controlling the disease in localized region with lower population and indicates that awareness, accurate information of local outbreak and the subsequent preventive measures are keys to fight the disease spread while keeping the economic activities on the move, of course with restrictions.
4. The unlock procedure can be easily made based on the quantification (estimating the b_0_ and c or B_0_ and C values for a particular region) of how Covid spread is controlled or inhibited in a region. This implies starting of communication between large regions of a city, between cities, between states and between countries. The regions with similar value or close values of disease inhibition or control parameters are in principal compatible and least risk of worsening the situation in the regions is expected when the movement restrictions between them are lifted.
5. Increasing awareness, fast communication, smart and dynamic administration, authentic data sharing and upkeep, coherent discipline and adherence to social distancing and hygiene and sanitization are the key for ensuring the disease control and inhibition and for preventing mortality.

## Data Availability

The active case data is obtained from https://www.worldometers.info/coronavirus/
which is open access data

https://www.worldometers.info/coronavirus/

## Data

The active case data is obtained from https://www.worldometers.info/coronavirus/

## Acknowledgement

The Authors are thankful to Dr. M.P. Singh, Theory & Simulations Lab, RRCAT for his useful comments on the manuscript. The Authors are highly grateful to Shri Rakesh Kaul, Associate Director, Materials Science Group & Head, Laser Materials Processing Division & Materials Engineering Section of RRCAT for his useful comments, suggestions and support.

